# Transcranial magnetic stimulation of dorsomedial prefrontal cortex for cocaine use disorder: A pilot study

**DOI:** 10.1101/2024.09.16.24313754

**Authors:** Heather E. Webber, L. Elliot Hong, João Quevedo, Michael F. Weaver, Joy M. Schmitz, Scott D. Lane

**Author notes:** Correspondence regarding this article can be sent to Heather E. Webber, 1941 East Rd., Behavioral and Biomedical Sciences Building, CNRA 1^st^ floor Houston, TX 77054.

## Abstract

Cocaine use disorder (CUD) is a difficult-to-treat condition with no FDA-approved medications. Recent work has turned to brain stimulation methods to help rectify hypofrontality and dopamine reward system changes often observed in individuals with CUD. Preliminary studies using transcranial magnetic stimulation (TMS) have demonstrated promising results, but there is room for optimization of the stimulation site, stimulation pattern, and identification of relevant biomarkers of TMS effects. The current pilot study aimed to test the feasibility, safety, and preliminary effects of a double-blind, sham-controlled, cross-over, acute design using intermittent theta burst stimulation to dorsomedial prefrontal cortex (dmPFC) on electroencephalogram (EEG) as intermediate outcome assessment in CUD patients. This small pilot enrolled five individuals with moderate-to-severe CUD for feasibility and proof-of-concept. Participants completed safety, psychometric, and EEG measures before and after receiving two sessions of active or sham TMS to dmPFC on two separate days. All five participants completed all the study tasks and found the TMS to be tolerable. The side effects were minimal and consistent with an acute TMS design. Visible changes were observed in the electrical activity of the brain during a monetary guessing task, while minimal changes in psychometric measures were observed. These results indicate the feasibility and safety of the current approach and suggest that dmPFC is a viable target for treating CUD. Future work should expand upon these findings in a randomized controlled clinical trial.

## Introduction

Cocaine use disorder (CUD) is an often unremitting condition that leads to negative health outcomes^1^. Despite well-known effects of chronic cocaine use on the dopamine reward system^2^, treating deficits in dopamine and other monoamine functioning with pharmacological agents has led only to small-to-moderate improvements^3^. There are currently no FDA-approved medications for the treatment of CUD. Alternative approaches that directly target neural circuits using brain stimulation may hold promise for treating CUD-related deficits.

Transcranial magnetic stimulation (TMS) is a form of brain stimulation that involves generating a magnetic field that passes through the skull by passing an electrical current through an insulated coil. Repetitive TMS (rTMS) can have excitatory or inhibitory effects on the brain area directly under the coil and associated networks. rTMS is an FDA-approved treatment for treatment resistant depression, obsessive compulsive disorder, and short-term smoking cessation^4,5^. Recent preliminary research has also shown promising results for the use of rTMS for the treatment of CUD^6–8^. It is well known that CUD is associated with changes in prefrontal functions including executive and emotional control, inhibition, decision-making, and processing of motivationally salient stimuli^9–13^. As a result, the majority of TMS studies in CUD thus far has aimed to increase the activity of the prefrontal cortex by applying excitatory stimulation to the left dorsolateral prefrontal cortex (dlPFC). Stimulation of dlPFC could also rectify changes in reward system networks as it projects to midbrain dopamine cells within the ventral tegmental area and substantia nigra^14^. Preliminary evidence has shown that multiple sessions of active rTMS of 5-15 Hz to dlPFC reduces self-reported craving, cocaine positive urines, and other unwanted symptoms^6,15–22^.

Despite considerable promise, there are addressable issues for optimizing treatment with TMS. The first issue involves identifying the optimal stimulation site. While the majority of rTMS studies have stimulated dlPFC to rectify the prefrontal hypoactivation commonly observed in CUD, other prefrontal structures involving emotional and reward functioning are also potential sites for TMS^10^, including dorsomedial PFC (dmPFC) and its connections to the anterior cingulate cortex (ACC). dmPFC is a promising target for TMS, as people who use cocaine show reduced dmPFC and ACC activity in response to pleasant emotional images/monetary outcomes^23–25^, and reduced functional connectivity of mesocorticolimbic circuits^26^. There are limited studies stimulating dmPFC in CUD, but recent evidence suggests that TMS to dmPFC could be beneficial in treating depressive symptoms^27–29^.

The next TMS parameter that needs to be optimized is the stimulation pattern. In addition to a traditional 10/15Hz rTMS protocol, intermittent theta burst stimulation (iTBS) has also shown early success in lowering cocaine use and craving^30^. iTBS protocols take about 3 minutes to complete compared to the 15 minutes it takes for a 15Hz protocol and have similar clinical effectiveness for treatment of depression^31^. Steele et al. (2019) reported iTBS to left dlPFC reduced cocaine use and craving^32^, while Sanna et al. (2019) reported no differences in 15 Hz stimulation compared to iTBS to the bilateral PFC on cocaine use^33^. Taken together, iTBS to either area could be useful in treating CUD.

The third issue involves identifying relevant targets and alternative end points for treatment studies. The majority of studies have used the subjective measure of cocaine craving as the main outcome^30,34^. While subjective measures are useful, they may not capture all the heterogeneity in TMS effects^6^. rTMS alters the electrical activity of the brain, while event-related potentials (ERPs) directly measure the electrical activity of the brain with millisecond temporal prescision^35^, making ERPs an ideal way to objectively assess rTMS effects. Two ERP components, Reward Positivity (RewP) and Late Positive Potential (LPP), might be relevant to rTMS effects in CUD, as they reflect overall reward sensitivity and motivated attention to drug cues, respectively^36–38^ and are altered in those with substance use disorders^39,40^. The RewP is thought to reflect a reward prediction error signal from midbrain dopamine neurons to the ACC^41–48^, while the LPP is thought to arise from a complex network of brain areas, including the visual cortex, amygdala, prefrontal cortex, and insula^49–51^. Our prior work has shown that the RewP and LPP are altered in individuals with substance use disorders and are associated with addiction-related constructs^52–57^. Further, these ERPs are sensitive to TMS manipulation^58–60^, but no study has assessed the effects of rTMS to dmPFC specifically or in a sample of individuals with CUD. Measuring both RewP and LPP will provide clarification on whether TMS alters overall reward functioning or a bias toward drug rewards compared to non-drug rewards.

This preliminary study aimed to pilot the feasibility of a double-blind, cross-over, sham-controlled, acute rTMS protocol in individuals with CUD (n = 5) in an outpatient research setting. The participants completed self-report and behavioral tasks and were administered an electroencephalogram (EEG) to measure the RewP and LPP components immediately before and after two successive iTBS sessions that targeted the dmPFC. Participants also competed these tasks with sham stimulation. Safety measures and completion rate were also collected to assess the safety and feasibility of the protocol in this population.

## Methods

### Overall Study Design

A double-blind, cross-over, sham-controlled, pre/post study design was used. Each participant completed two test days: 1) Active iTBS to dmPFC and 2) Sham iTBS to dmPFC. The order (dmPFC or sham) of test days was counterbalanced across participants and double-blind. Participants completed screening and baseline measures prior to undergoing the two-day within-subjects pre/post rTMS design. On each day, participants completed self-report, behavioral and EEG tasks immediately before and after receiving two iTBS sessions. All procedures were approved by the local IRB. This study was registered on ClinicalTrials.gov (NCT05631548).

### Participants

Five participants were enrolled into the pilot study at a research clinic in the greater Houston area. Eligible participants were between the ages of 18 and 65 and met DSM-5 criteria for moderate-to-severe CUD. Participants were excluded if they met moderate or severe criteria for substances other than cocaine, cannabis, or nicotine, had an unstable psychiatric disorder, had a medical condition contraindicated to TMS (e.g., medical implants, history of seizure or seizure disorder, medications lowering the seizure threshold, neurological conditions, moderate-to-severe heart disease, brain surgery, stroke), if they were pregnant, or had a head injury with loss of consciousness. Screening criteria were assessed with the Structured Clinical Interview for DSM-5 (SCID)^61^, Colombia Suicide Severity Rating Scale (C-SSRS)^62^, Assault & Homicidal Danger Assessment Tool^63^, urine drug screening, breath alcohol testing, urine pregnancy tests, and the TMS safety screen^64^. Participants received monetary compensation for completing the screening and the study tasks.

### TMS Protocol

A trained nurse and/or research assistant administered the TMS. Prior to each TMS session, we collected the following information to ensure the safety of the participants: hours of sleep, urine drug screen, Timeline Followback^65^, and breathalyzer. If participants slept less than their normal amount, tested positive for or reported using any substances other than cocaine, marijuana, or alcohol, or blew above .000 on the breathalyzer, then their appointment was rescheduled. TMS was delivered with a MagVenture Mag Pro R30 with the Cool-B70 A/P coil with active liquid cooling and active/sham sides (Farum, Denmark). A member of the clinic who was not involved in the study was the only person aware of the randomization results and provided the randomization code for the system to ensure double blinding. The first session began with the acquisition of the resting motor threshold (rMT; lowest stimulus intensity that elicits a visible thumb twitch on 50% of the trials) on the contralateral hand. Two electrodes were affixed to the forehead on both days for blinding and sham purposes. dmPFC was targeted by measuring approximately 25% of the nasion-inion distance consistent with prior studies^28,66^. iTBS (triplet 50 Hz bursts, repeated at 5 Hz, 2 sec on and 8 sec off; 600 pulses per session) were delivered at 80% of the rMT and lasted ∼3 minutes, consistent with current EEG-iTBS single day protocols^58,67^. The intensity was lowered if the participant could not tolerate the stimulation. Each participant received two iTBS sessions with a 15-20 minute interval between sessions in order to boost acute rTMS effects^68^.

### Measures

#### Participant Characteristics

Participants completed the Addiction Severity Index-Lite^69^ during their screening to measure their demographic information and cocaine use patterns, including years of use and days of use in the past 30 days.

#### Feasibility and Safety Measures

Completion rate/study adherence was assessed to measure the feasibility of the protocol in this population and setting. *Cognitive Function* was measured with the Montreal Cognitive Assessment (MoCA)^70^ which includes multiple cognitive domains (attention, concentration, executive functions, memory, language, visuospatial skills, abstraction, calculation, and orientation). The MoCA was administered before and after the TMS protocol to ensure that the TMS protocol did not impair cognitive functioning. Alternative equivalent forms were used for the pre- and post-tests. *Side Effects* were measured with an AE form where participants marked off if they experienced the side effect and to what extent (none, mild, moderate, severe). *Pain* was also assessed on a scale of 0-10. The AE form and pain were assessed before and after the TMS protocol. Participants were verbally asked about their experience with the TMS sessions and how they felt when they went home at the following session. Effectiveness of the *Blinding and Sham* procedure was measured by asking the participants if they thought they received the real TMS or fake TMS after each session.

#### EEG Acquisition & Tasks

During the tasks described below, EEG was collected using a 64-channel actiCAP electrode cap, amplified with BrainAmp MR and digitized using Brain Vision Recorder (Brain Products, Munich). Impedances were maintained below 50 k(OHM). The sampling rate was 500 Hz and data were filtered online (0.1 Hz high-pass/100 Hz low-pass). <u>The</u> <u>Doors Task</u>^59^ was used to elicit the RewP component, representing reward sensitivity^47,60^. The task is a guessing game, where participants guess which door contains a reward behind it. After selecting a door, the participants are notified if they found the prize by a green arrow pointing up or if they did not find the prize by a red arrow pointing down. Unknown to the participants, winning and losing outcomes are presented 50% of the time in a random order. The RewP is measured in response to the outcome (up or down arrow). <u>The Picture Viewing Task</u> was used to elicit the LPP, reflecting the motivational salience of a stimulus. During this task, participants are asked to view a slideshow of images including pleasant, unpleasant, neutral, and cocaine-related images^61^. These EEG Tasks were administered before and after the TMS protocol.

#### Psychometric Measures

Self-reported anhedonia (Snaith Hamilton Pleasure Scale; SHAPS^71^) was assessed as a secondary measure of reward sensitivity in addition to the RewP. Cocaine craving was measured using the Minnesota Cocaine Craving Scale (MCCS)^72^. Only the craving intensity subscale was used. These self-report measures were administered before and after the TMS protocol.

### Data Analysis Plan

#### EEG Analysis

EEG data were analyzed using Brain Vision Analyzer (Brain Products, Germany). Data were filtered (.01-40 Hz) offline and re-referenced to the average reference. For the Doors Task, the data were segmented into 1000ms segments, 200ms before and 800ms after the onset of the feedback arrow. For the Picture Viewing Task, the data were segmented into 1200ms segments, 200ms before and 1000ms after the onset of the images. Blinks were detected and corrected using Brain Vision Analyzer’s Ocular Correction Independent Components Analysis tool. Artifacts were detected using the following criteria: 1) 10uV/ms maximal allowed voltage step, 2) 70uV maximal allowed difference in a 50uV interval, 3) +/-50uV maximum/minimum amplitude, and 4) 0.5uV lowest allowed activity in 100ms interval.

Channels were marked bad if 40% of the segments had an artifact and were interpolated with the surrounding 8 channels. Segments were removed if 10% or more of the channels were marked bad. Cleaned segments were then sorted and averaged by condition (wins/losses for RewP; pleasant, unpleasant, cocaine, and neutral for LPP) and by session (pre/post sham and active TMS). Grand average ERPs were visually inspected for differences within the 200-350ms time range at electrode FCz for the RewP and within the 400-800ms time range over the central-parietal electrodes (Cz, C1, C2, CPz, CP1, CP2) for the LPP.

#### Hypothesis Testing

This pilot study was focused on evaluating the feasibility of the current approach with respect to recruitment, retention, assessment procedures, implementation, and safety. For that reason, no statistical tests were performed. For the preliminary outcome measures, participant-level means are presented and visually inspected for differences between conditions, except for the ERPs. Because individual ERPs are often noisy and difficult to interpret, all 5 participants were averaged together for visual inspection. Grand average means for all outcomes are also presented to evaluate preliminary changes observed.

## Results

### Feasibility and Safety

Participant characteristics are displayed in *Table 1*. Ten people were approached and participated in the initial screening. Five participants were excluded for the following reasons: 1) three had a family history of seizures or seizure disorder, 2) one met criteria for an alcohol use disorder, and 3) one had uncontrolled high blood pressure. Five participants met criteria for the study and were enrolled and randomized. All five participants had a 100% completion rate, attending the screening and both study visits. Additionally, all participants met the TMS safety criteria at the start of the TMS day and did not have to reschedule their appointment.

**Table 1.** Participant Characteristics.

| Characteristic | M(SD)/% (n = 5) |
| --- | --- |
| Age | 45.80 (12.48) |
| Sex | 100% Male |
| Race | 60% Black/African American |
| Ethnicity | 80% Not Hispanic |
| Education | 80% High School Degree |
| 30 Days Cocaine Use | 13.40 (10.97) |
| Lifetime Cocaine Use | 19.00 (12.21) |

Four participants found the TMS to be tolerable at 80% rMT. One participant asked for the intensity to be turned down for both the active and sham sessions. TMS machine intensity was as follows by participant: P1 = 45%; P2 = 40%; P3 = 50%; P4 = 62%; P5 = 62%. Responses to the AE form are displayed in *Table 2*. To summarize, no severe side effects were reported post-TMS. Mild neck pain, mild muscle spasms, and mild lightheadedness were reported by one participant each on the active TMS session day. Mild-to-moderate scalp discomfort was reported by 3/5 participants and mild-to-moderate headache was reported by 2/5 participants. On the sham day, one participant reported mild tingling; one participant reported mild scalp discomfort; and one reported mild lightheadedness after the TMS protocol. For overall pain, four participants reported minimal pain for both sham and active days. One participant reported an increase in pain pre/post from 1/9 on the active day and 1/5 on the sham day (*Figure 1G/1H*). Two participants reported a lingering headache that was alleviated with over the counter analgesics.

**Figure 1.**
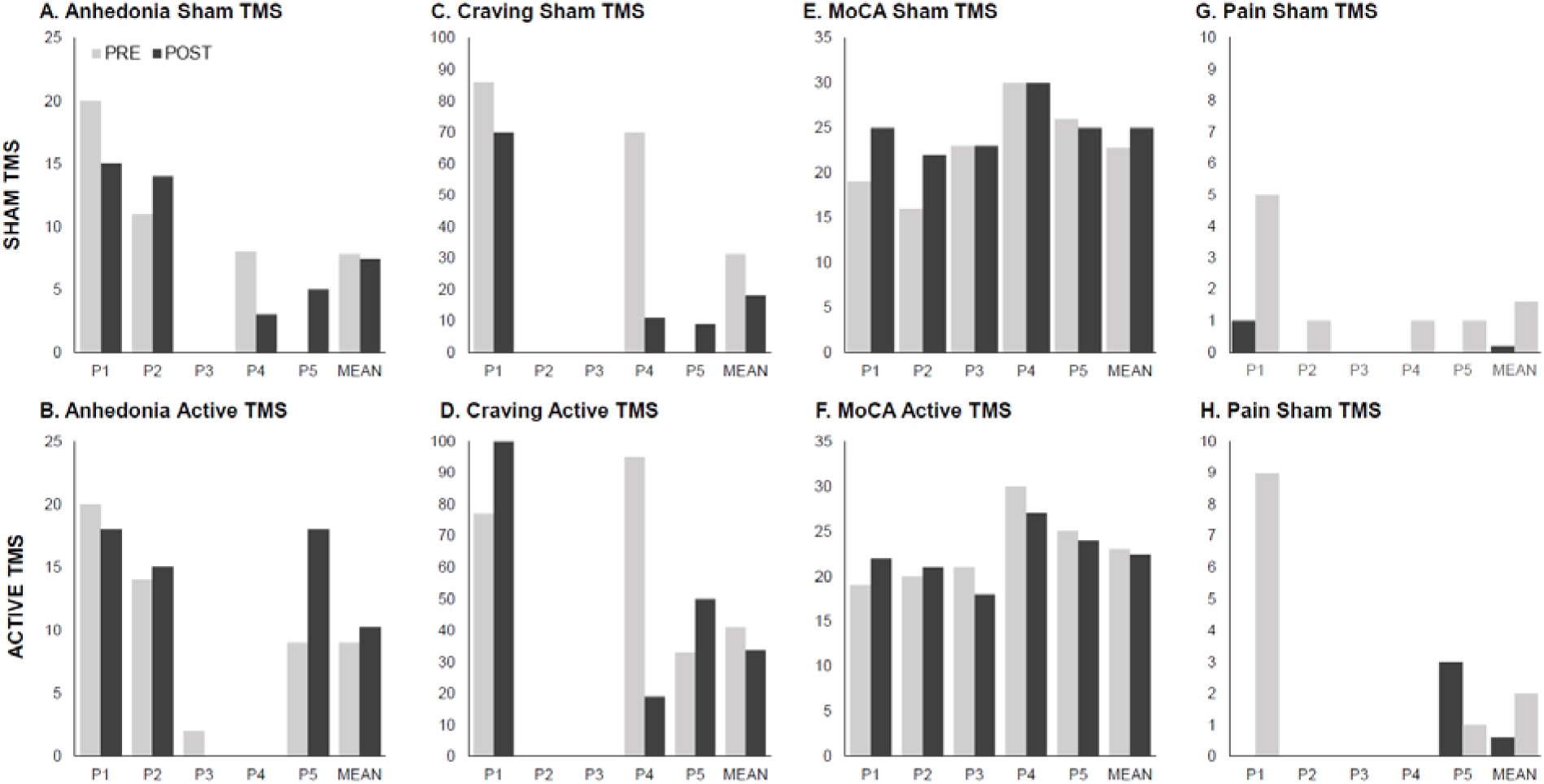
Participant and mean level results for psychometric and safety outcomes.

**Table 2.** Side Effects Questionnaire by Participant.

|  | Pre-Active TMS |  |  |  |  | Post-Active TMS |  |  |  |  |
| --- | --- | --- | --- | --- | --- | --- | --- | --- | --- | --- |
| Participants (1-5) | P1 | P2 | P3 | P4 | P5 | P1 | P2 | P3 | P4 | P5 |
| **Side Effects** |  |  |  |  |  |  |  |  |  |  |
| Scalp Discomfort | 0 | 0 | 1 | 0 | 1 | 2 | 0 | 0 | 1 | 1 |
| Neck Pain | 0 | 0 | 0 | 0 | 0 | 1 | 0 | 0 | 0 | 0 |
| Headache | 0 | 0 | 0 | 0 | 0 | 2 | 0 | 0 | 0 | 1 |
| Tingling | 0 | 0 | 0 | 0 | 0 | 0 | 0 | 0 | 0 | 0 |
| Muscle Spasm | 0 | 0 | 0 | 0 | 0 | 1 | 0 | 0 | 0 | 0 |
| Twitching Eye | 0 | 0 | 0 | 0 | 0 | 0 | 0 | 0 | 0 | 0 |
| Lightheaded | 0 | 0 | 0 | 0 | 0 | 1 | 0 | 0 | 0 | 0 |
|  | Pre-Sham TMS |  |  |  |  | Post-Sham TMS |  |  |  |  |
| Participants (1-5) | P1 | P2 | P3 | P4 | P5 | P1 | P2 | P3 | P4 | P5 |
| **Side Effects** |  |  |  |  |  |  |  |  |  |  |
| Scalp Discomfort | 0 | 0 | 0 | 0 | 2 | 0 | 0 | 0 | 1 | 0 |
| Neck Pain | 0 | 0 | 0 | 0 | 0 | 0 | 0 | 0 | 0 | 0 |
| Headache | 0 | 0 | 0 | 0 | 3 | 0 | 0 | 0 | 0 | 0 |
| Tingling | 0 | 0 | 0 | 0 | 3 | 1 | 0 | 0 | 0 | 0 |
| Muscle Spasm | 0 | 0 | 0 | 0 | 1 | 0 | 0 | 0 | 0 | 0 |
| Twitching Eye | 0 | 0 | 0 | 0 | 1 | 0 | 0 | 0 | 0 | 0 |
| Lightheaded | 0 | 0 | 0 | 0 | 2 | 1 | 0 | 0 | 0 | 0 |

*Note.* 0=None, 1=Mild, 2=Moderate, 3=Severe

Results regarding the MoCA are displayed in *Figure 1E/1F*. Overall, there were minute changes in the MoCA pre/post sham and active TMS for all participants. In terms of the success of the sham procedure, participants were always told that they would receive one day active and one day sham, but three participants guessed that they had “real” TMS on both days, one guessed the days correctly, and one guessed incorrectly.

### EEG Measures

Grand average RewP amplitudes are displayed in *Figure 2*. Minimal to no differences were observed pre/post sham TMS for wins or for losses. The only visual change in the RewP amplitude was to losses on the active TMS day. Compared to baseline, post-active TMS amplitude to loss was larger (more negative).

**Figure 2.**
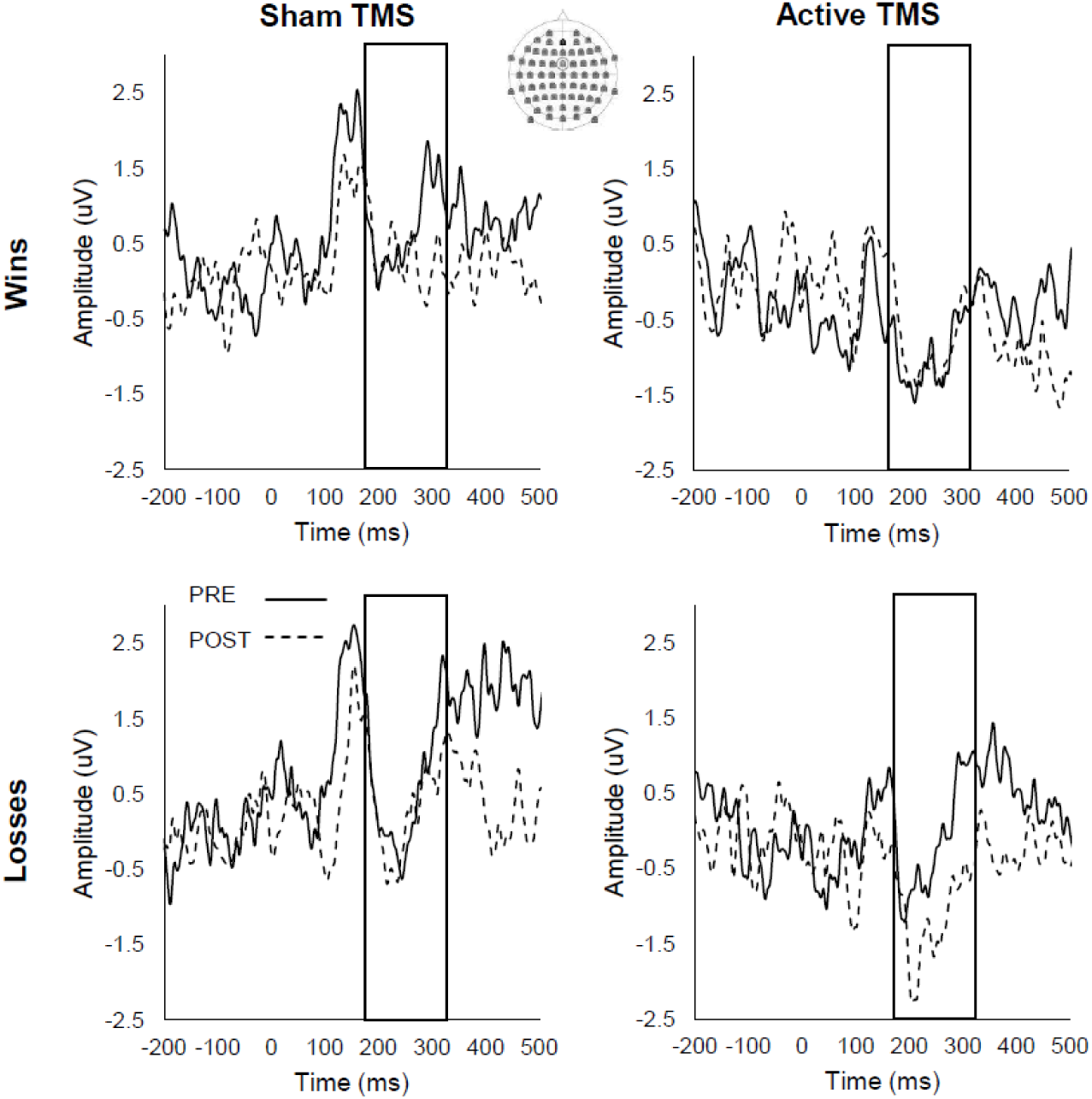
Grand mean RewP amplitudes. Top left: RewP to wins pre/post sham stimulation. Bottom left: RewP to losses pre/post sham stimulation. Top right: RewP to wins pre/post active stimulation. Bottom right: RewP to losses pre/post active stimulation. Amplitudes are displayed at electrode FCz.

Grand average LPP waveforms for the LPP are displayed in *Figure 3*. Visually, small increases in the LPP to pleasant and neutral were observed on the sham day, while a small increase to cocaine was observed on the active TMS day.

**Figure 3.**
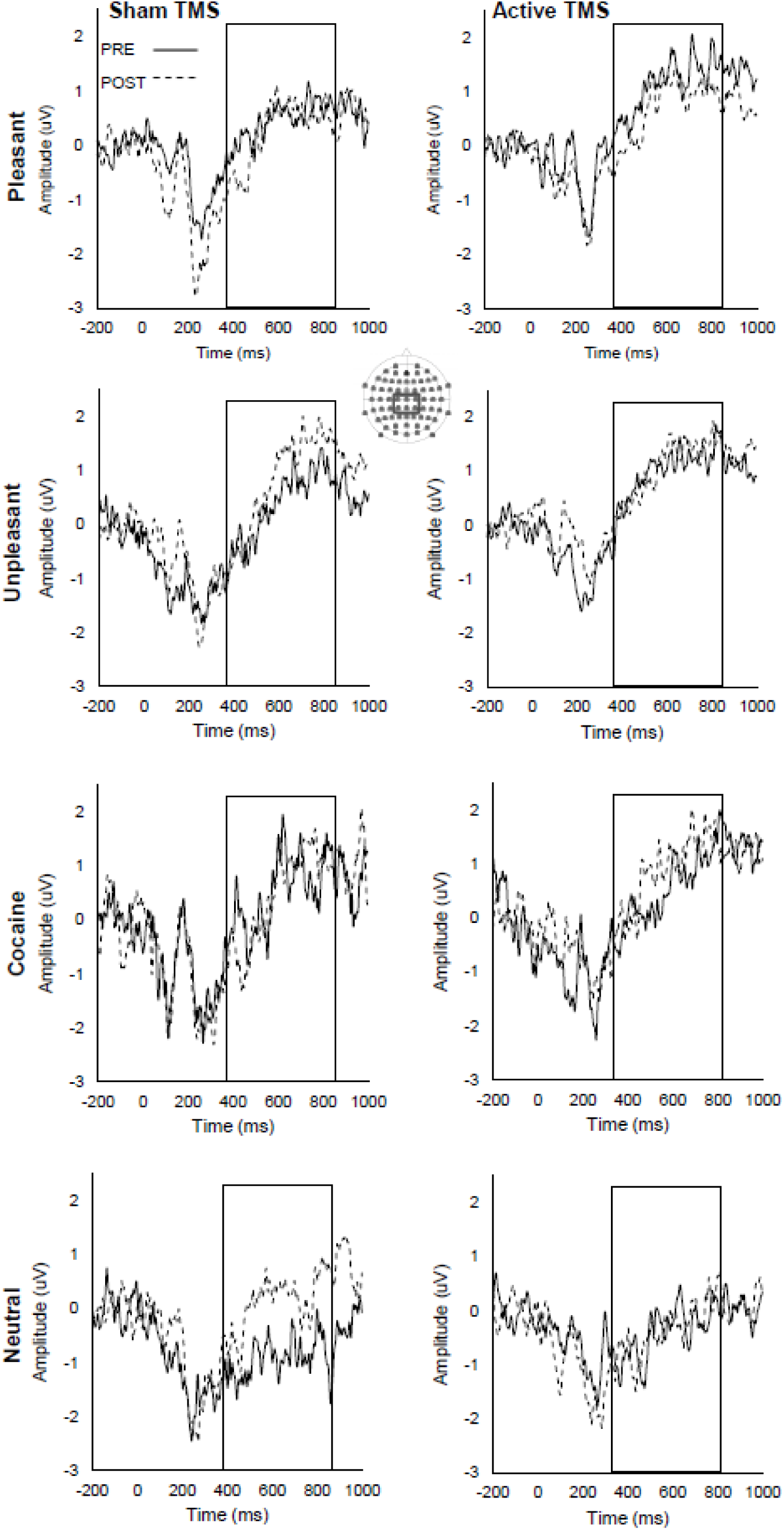
Grand mean LPP amplitudes. Left column: LPP amplitudes pre/post sham stimulation. Right column: LPP amplitudes pre/post active stimulation. Rows from top to bottom: LPP to pleasant, unpleasant, cocaine, and neutral images. Amplitudes are displayed as an average of the highlighted electrodes.

### Psychometric Measures

Overall, TMS did not appear to have a consistent effect on self-reported anhedonia across participants. One participant had a moderate increase in anhedonia following active TMS; otherwise, any changes were minimal. Craving was also inconsistent. Two participants did not report any craving pre or post TMS. Two had increases in craving and one had a very large decrease in craving following active TMS.

## Discussion

The present study aimed to test the feasibility and safety of a TMS protocol that stimulated dmPFC using iTBS in a sample of individuals with CUD. Overall, the protocol was found to be safe and tolerable for the participants. The protocol was also feasible, as demonstrated by the 100% completion rate of all study tasks. While one participant was particularly sensitive to the stimulation (both sham and active), this did not lead to their discontinuation from the study. Below, we discuss how this preliminary study can be expanded upon to aid in the optimization of TMS treatments for CUD.

First, the success of this preliminary study highlights the viability of utilizing TMS, specifically iTBS to dmPFC, in this population. iTBS is known to cost less in a clinical setting^73^ and has been found to be non-inferior to the standard 10Hz in terms of effectiveness for treatment-resistant depression^31,74^. These studies also found similar side effects between the two types of stimulation, with headaches being the most common adverse event. iTBS has been investigated in patients with CUD in two studies, to our knowledge. In an open label feasibility study, Steele et al., 2019 found multiple sessions of iTBS to dlPFC to be tolerable, with headache the most common adverse event^32^. Sanna et al., 2019 used the deep H4 coil that stimulates insula and bilateral prefrontal cortex in patients with CUD and also found that some participants reported headaches, dizziness, and sleepiness^33^. The location of the coil can affect tolerability and side effects of TMS; however, we found that stimulation to dmPFC here was similar to the other studies stimulating dlPFC and bilateral PFC. These results indicate that the coil location utilized in the present study could be safely investigated further in future studies. Further, as the protocol was tolerable, a higher % rMT may be investigated in future studies.

In addition to assessing the feasibility and safety of this TMS protocol, we also collected preliminary data on EEG and psychometric outcomes. EEG is particularly well-suited to assess changes in the brain associated with TMS, as it directly measures the electrical activity of the brain. Our results supported the use of EEG to measure TMS outcomes compared to the psychometric measures. Mean level changes were observed for both the RewP and LPP, but minimal changes were observed for self-reported craving and anhedonia. Inclusion of EEG in addition to the self-report measures might be useful in future trials aiming to assess longitudinal effects of TMS treatment.

A few studies have investigated the effects of TMS on the RewP and the LPP. One stimulated the rostromedial prefrontal cortex and found an increase in the RewP specifically to wins^75^. Another stimulated dlPFC in a sample of people who used substances and found that stimulation of dlPFC enhanced the RewP^76^. TMS to dlPFC also altered the RewP specifically to cigarette rewards compared to monetary rewards^59^. In the present study, we stimulated dmPFC in a sample of adults with CUD. While we only collected 5 participants, our results are consistent with others that indicate the RewP is sensitive to TMS. However, our results were specific to losses rather than wins. Future studies with a well-powered sample size will be useful in determining if these differences are due to the small sample size here, differing prefrontal stimulation site, stimulation pattern, population under study, or another factor. A few studies have also investigated the effects of TMS on the LPP. Stimulation to the left ventrolateral PFC enhanced the LPP to reappraisal stimuli^77,78^. Alternatively, stimulation to right ventrolateral PFC reduced the LPP to reappraisal stimuli specifically in a social pain condition^60^. To our knowledge, no study has investigated the effects of dmPFC stimulation on the LPP. Our preliminary results indicated a potential slight increase in the LPP to cocaine images post/active TMS. However, the LPP was also enhanced post-sham TMS to other emotional images.

Therefore, the results may not be specific to the TMS procedure, but to other events that occurred during the experiment.

In conclusion, the present TMS protocol was safe and feasible, and preliminary evidence supports the viability of using EEG to measure TMS effects in a sample of participants with CUD. These preliminary data are not expected to provide a meaningful estimate that would generalize beyond the small sample observed here. By providing evidence for feasibility, the present study will inform subsequent studies with formal hypothesis testing. This approach to pilot studies is explicitly in line with current literature on trial design^79^. Future work should perform these procedures in a larger sample. Additional future directions include comparing dmPFC to other PFC locations like dlPFC, comparing EEG outcomes to reward-related behavioral outcomes, increasing the intensity of the stimulation, increasing the number of sessions, and assessing TMS effects on brain circuits by adding fMRI as an outcome. Work along these lines will facilitate the optimization of parameters needed to enhance TMS treatment for CUD and other substance use disorders.

## Author Notes

### Funding

This work was supported by the UTHealth Houston Career Development and Research Excellence (CaDRE) Program in Psychiatry. The work by HEW was funded in-part by NIDA K01DA058765.

## Data Availability

All data produced in the present study are available upon reasonable request to the authors.

## Acknowledgements

The authors would like to thank Vincent Dang, Lanelle Ochiam, and Kimberly Boyette for their assistance in collecting the data. The John S. Dunn Distinguished Professorship funds the Center for Interventional Psychiatry research operation.

## Conflicts of Interest

JQ has a clinical research support relationship with LivaNova; is a member of the speaker bureau with Myriad Neuroscience and AbbVie; is a consultant for EMS, Libbs, and Eurofarma; is a stockholder at Instituto de Neurociencias Dr. Joao Quevedo; and receives copyrights from Artmed Editora, Artmed Panamericana, and Elsevier/Academic Press.

## Notes

### Clinical Trial

NCT05631548

### Funding Statement

This study was funded by the UTHealth Houston Career Development and Research Excellence (CaDRE) Program in Psychiatry. The work by HEW was funded in-part by NIDA K01DA058765.

### Author Declarations

The Committee for the Protection of Human Subjects of the University of Texas Health Science Center at Houston gave ethical approval of this work.

